# Baseline markers of treatment response to vagus nerve stimulation in difficult-to-treat depression: a retrospective data analysis

**DOI:** 10.1101/2025.08.31.25334709

**Authors:** Michael C. Treiber, Bernhard T. Baune, Anna Gramser, Johan Saelens, Natalia Grüll, Karl Rössler, Klaus Novak, Aicha Bouzouina, Sharmili Edwin Thanarajah, Christine Reif-Leonhard, Erhan Kavakbasi, Christoph Kraus

## Abstract

**Background:** Vagus nerve stimulation (VNS) is an FDA-approved invasive neuromodulatory treatment for Difficult-to-treat-Depression (DTD). However, approximately 50% of patients do not respond sufficiently to VNS. A previous study found that the baseline corrected QT (QTc) interval correlates with later VNS treatment response. We aimed to replicate this finding and further explored physiological and blood-based predictors of VNS response.

**Methods:** Data of 53 patients treated with VNS were pooled between three VNS centres. All available QTc data and blood-based parameters were used. We conducted a regression analysis with baseline QTc length and Montgomery-Åsperg Depression Rating Scale (MADRS) changes after six- and 12- months of VNS. For analysis of blood-based variables, we used exploratory correlation analysis with a stepwise forward regression approach.

**Results:** After correction for sex, age and body mass index (BMI) and medication, baseline QTc intervals did not correlate with MADRS change at either follow-up. Exploratory regression analysis identified four factors associated with MADRS reduction after six-months of VNS: lower absolute neutrophil count (ANC), BMI, younger age, and higher low-density lipoprotein (LDL) levels. Only absolute neutrophil count correlated with MADRS change after 12-months VNS.

**Discussion:** With a larger sample and relevant covariates, we did not replicate a previous finding that baseline QTc interval correlates with later MADRS change upon antidepressant VNS. However, in exploratory analyses, blood-based markers indicated links between MADRS change and ANC, BMI, age, and serum LDL. These findings need to be verified in prospective, controlled studies to facilitate optimal prescription of antidepressant VNS.

## Background

Major depressive disorder (MDD) is a heterogeneous and complex disease, which impacts on all forms of social and daily functioning and exhibits excess mortality including increased suicidal risk (Marx et al., 2023). An early remission is critical for improved long-term outcomes (Rush et al., 2006). Approximately one fourth of individuals with MDD do not sufficiently remit after multiple treatment trials. Difficult-to-treat depression (DTD) mischaracterised by increased morbidity and mortality, higher rates of relapse and suicide rates, and increased rates of hospitalisations with a higher burden for mental and physical health services costs (Amos et al., 2018; Bergfeld et al., 2018; McAllister-Williams et al., 2020; Rush et al., 2006).

Vagus nerve stimulation (VNS) is an implantable neuromodulatory technique for long-term treatment of DTD, either in unipolar or bipolar affective disorders. Clinically, VNS is an adjunctive treatment for patients with DTD who did not achieve or sustain response to a minimum of four antidepressant treatment trials including psychopharmacological treatments, psychotherapies and electroconvulsive therapy (Demyttenaere et al., 2024). An open label registry study reported a reduction of depressive symptomatology of about 50% in approximately one third of patients within the first year of VNS adjunctive to treatment as usual (Aaronson et al., 2017). Recently, a randomized, one year sham-controlled trial in the U.S.A. investigated the effects of active VNS compared to sham-treatment (RECOVER-trial). Conway et al. (2025) reported a very high disease burden with extensive and unsuccessful exposure to antidepressant treatment in these patients (Conway, Aaronson, Sackeim, Duffy, et al., 2024). The study did not achieve the primary endpoint, which was the difference in depressive symptom reduction as assessed by the MADRS over a period of 12 months (Conway, Aaronson, Sackeim, George, et al., 2024). Still, VNS significantly improved secondary outcomes such as quality of life, psychosocial functioning, and depression self-rating scores (Conway, Aaronson, Sackeim, George, et al., 2024; Rush et al., 2024). In Europe, the RESTORE-LIFE study is an ongoing open-label, prospective, multi-center, naturalistic, observational study evaluating the long-term effectiveness and safety of adjunctive VNS in individuals with DTD. In this study, so far, older age and increased symptom severity have been related to increased symptom reduction after 12 months of VNS treatment (Kavakbasi et al., 2025).

Given that VNS requires surgery and generator and electrodes are hard to explant, estimates of future response would constitute a substantial improvement towards personalized indication. Optimally, VNS would be preferentially prescribed to patients with higher chances of response to VNS. In that regard, the corrected QT interval (QTc), an indirect marker of the sympathovagal balance, correlated with treatment response to antidepressant VNS (Longpré-Poirier et al., 2020). The authors investigated the association between baseline QTc intervals and changes in Hamilton Depression Rating Scale (HAM-D) in 19 patients suffering from MDD. In this study, longer baseline QTc intervals prior to VNS correlated with greater reductions in HAM-D scores 12 and 24 months after VNS. However, relevant covariates for QTc such as sex were not considered (Jayanthi et al., 2021). To date, no other physiological predictive markers have been reported. Taken together, little is known about which baseline variables correlate with long-term response to VNS treatment. Hence, the primary aim of this study was to replicate previous correlations between baseline QTc interval and reduction of depressive scores in an independent sample. As secondary aim, we explored other available baseline variables obtained prior to VNS available at our study centre. The overall future goal of this line of evidence is to improve selection of patients potentially benefitting from VNS treatment.

## Methods

### Study sample

Patients included in this retrospective data analysis were taking part in the ongoing RESTORE-LIFE study. A detailed description of the RESTORE study protocol and procedures has been published (Young et al., 2020). In this study, we included patients recruited at three sites (Frankfurt am Main, Münster, Vienna). Individuals 18 years of age or older were eligible if they met criteria for a primary diagnosis of recurrent or chronic major depressive episode (unipolar or bipolar) according to the *Diagnostic and Statistical Manual of Mental Disorders* (5^th^ edition), and had not achieved treatment response to an adequate number of antidepressant treatment trials. Diagnosis was made based on the Mini-International Neuropsychiatric Interview (MINI). Exclusion criteria were (1) severe substance use disorder, (2) predominant psychosis, (3) severe personality disorder, and (4) mental retardation (Young et al. 2020). The study was conducted in accordance with the Declaration of Helsinki. All study procedures were approved by the Ethics Committees of the Medical Universities of Vienna, Münster and Frankfurt am Main.

### VNS treatment

Following a psychiatrist’s decision to initiate VNS, eligibility was evaluated, and written informed consent was obtained. The baseline assessment was conducted within six weeks to one week prior to VNS implantation performed at the respective department of neurosurgery, trained psychiatrists conducted stimulation titration. Output current, pulse width, frequency, ON/OFF times were set during the titration visits. Taking safety and tolerability into account, the output current was increased in steps of 0.25mA, with the possibility of several steps of increase in one titration visit. Default frequency and pulse width was set at 20Hz, and 250msec, respectively, and could be adjusted by clinicians’ choice. For further information on dosing in this sample we refer to a previous publication (Kavakbasi et al., 2025). In default mode ON/OFF-time was set at 30sec, and 5min, respectively. To increase the duty cycle of VNS, the OFF-time could be decreased. Antidepressant treatments including psychotherapy, pharmacotherapy and neuromodulation (electroconvulsive treatment/transcranial magnetic stimulation) were continued according to clinical judgement.

### Measurements of Depressive symptoms

The MADRS was performed at baseline and every three months by trained raters, who were aware of the details of the trial. Remission was *a priori* defined as a total score ≤ 9. Response was a prior defined as remission or a symptom reduction of 50% (Rush et al., 2024).

### Electrocardiogram measurements

As a part of routine procedures, we retrospectively collected 12-lead standard electrocardiograms (ECG) recorded on a CardiofaxM ECG-1350 (Nihon Kohden), ChiroMed MAC 1600 (GE Healthcare), or Hellige Cardiognost EK 56 (CAmed Medical System) at a speed of 25mm/s and gain of 10mm/mV prior to neurosurgical implantation of the VNS-device. For each patient, a 12-lead standard ECG was recorded after laying down for 15min in a supine, relaxed position. Patients were instructed to breathe normally, and to avoid talking or moving. Heart rate was calculated from the RR-Interval. To obtain QTc-mean, QT-intervals were corrected for heart rate for the entire ECG using Bazett’s formula (QTc = QT/√RR interval), which is the most widely used formula in clinical and research practises (C Bazett, 1997). Following the American Heart Association criteria QTc prolongation was defined as >460ms in women and >450ms in men (Giudicessi et al., 2019). All ECG-recordings were reviewed by two experienced physicians.

### Blood-based measurements

For this study, blood-based measures were available only from the site in Vienna, thus we report explorative results from 22 patients. As part of routine procedures before surgery, venous blood samples were obtained from patients between 7:00 am and 8:00 am the day prior to surgical procedures. Patients were instructed to overnight fasting and tobacco abstinence for more than 12hours prior to blood sample collection. Blood samples were processed immediately to the laboratory for analyses of white blood cells, i.e., eosinophils, neutrophils, lymphocytes, monocytes, platelets, platelet distribution width, mean platelet volume and derived ratios, as well as metabolic parameters, such as fasting blood glucose, total cholesterol (CHOL), serum triglyceride (TG; reference range 50-200mg/dL), serum high-density-lipoprotein (HDL; reference range for standard risk 45-65mg/dL for women, 35-55mg/dL for men), serum low-density-lipoprotein (LDL; desirable for individuals without cardiovascular risk <150mg/dL). We further calculated inflammatory parameters, such as neutrophil to lymphocyte ratio (NLR), platelet-lymphocyte ratio (PLR), NLR and monocyte-lymphocyte ratio (MLR), and the systemic inflammatory index (SII = neutrophil count x platelet count / lymphocyte count (Šagud et al., 2023)).

### Anthropometric measurements

Anthropometric assessments included height, weight, and body mass index (BMI). Height was recorded to the nearest of 0.1 cm. For weight, patients were asked to remove their shoes, any heavy clothing (sweaters, jackets, or coats), and empty their pockets. Weight was measured to the nearest 0.1 kg. BMI was calculated as a ratio between weight and height (kg/m^2^).

### Statistical analysis

All statistical analyses were conducted with R statistical software version 4.3.2. Normal distribution was evaluated using Shapiro-Wilk test. The results indicated no significant deviation from normality (p > .05). We performed a Pearson correlation analysis to evaluate the association between MADRS score reduction and QTc length to relate our data to the previously published study by Longpré-Poirier et al., who included 19 patients in their study. To account for the effect of psychotropic medication on QTc length, we calculated a risk-score based on previous methods (Vandael et al., 2017). Accordingly, we coded a binary variable based on medication (1) with potential for QTc prolongation (e.g., quetiapine, tricyclics etc.) and (2) OTc neutral medication. We then calculated a multiple regression using QTc and MADRS score reductions after six- and 12-months VNS adjusting for age, BMI, sex, QTc prolongation risk-score and MADRS baseline scores. In accordance with previous studies, we chose the 12-months outcome, expecting the largest reductions in MADRS scores compared to baseline (Conway, Aaronson, Sackeim, George, et al., 2024; Kavakbasi et al., 2025; Kavakbasi & Baune, 2024; Lespérance et al., 2024; Longpré-Poirier et al., 2020; Rush et al., 2024). In addition, we chose six-months follow up in order to evaluate mid-term outcome, which is a standard time point to assess response to VNS (Kavakbasi & Baune, 2024).

To explore additional baseline variables correlating with later treatment response we conducted an exploratory analysis testing associations between MADRS score changes between six- and 12-months follow-ups and available baseline data. Thus, we used baseline inflammatory markers, metabolic markers and BMI. For the exploratory analysis we first performed Pearson correlation analyses to assess univariable associations between MADRS score reduction and: age, sex, BMI, MADRS baseline score, absolute neutrophil count (ANC), lymphocyte count, monocyte count, eosinophil count, basophil count, platelet count, NLR, MLR, PLR, SII, TG, HDL, LDL, CHOL, GLU. We considered a p-value < .05 as statistically significant. Since this analysis was exploratory, we did adjust for multiple comparisons. To account for intercorrelations between predictive markers and to further refine exploratory analyses, we than conducted stepwise multiple linear regression models with a forward selection approach. We used the change in MADRS scores as the dependent variable, and the following variables and independent variables: sex, age, ANC, lymphocyte count, monocyte count, eosinophil count, basophil count, platelet count, NLR, MLR, PLR, SII, TG, HDL, LDL, CHOL, GLU. We considered a p-value < .05 as statistically significant. The same strategy was replicated to explore associations between MADRS-score changes and baseline biomarkers at six- and 12-months follow-up.

## Results

### Sociodemographic and clinical characteristics

For this study 53 patients with DTD who received VNS at three study sites (Münster, Frankfurt, Vienna) were treated with VNS at all three sites. Baseline ECG and BMI data were unavailable for five patients; therefore, 48 patients constituted the primary analysis cohort. Sociodemographic and clinical characteristics are summarized in table 1. The mean age was 50.7 years (±11.7), 34 (70.8%) were female. Also, follow-up data availability varied across time points. Accordingly, we included ECGs from 45 patients for the six-months follow-ups and ECGs from 42 patients for the 12-months follow-up.

**Table 1.** Sociodemographic and clinical characteristics of the total sample (N = 48)

|  |  |
| --- | --- |
| Mean age at time of implantation (SD), years | 50.7 (11.7) |
| Female, % | 70.8 % |
| BMI | 30.1 (6.9) |
| BMI 18-25 | 12 (25%) |
| BMI 26-30 | 13 (27%) |
| BMI 30-35 | 23 (48%) |
| HR | 76.9 (13.4) |
| QTc | 428ms (26.8ms) |
| QTc female | 428ms (±27.2ms) |
| QTc male | 432ms (±23.8ms) |
| Baseline MADRS | 30.1 (8.9) |
| 3 months MADRS | 22.3 (8.6) |
| 6 months MADRS | 20.5 (10.1) |
| 9 months MADRS | 20.7 (9.9) |
| 12 months MADRS | 19.1 (9.0) |
| 6 months response rate, % | 32.7 % (16 out of 45) |
| 12 months response rate, % | 37.0 % (17 out of 42) |

*Note: BMI = body mass index; HR = heart rate; MADRS = Montgomery-Åsberg Depression Rating Scale; QTc = Bazett corrected QT interval; numbers in () represent standard deviation*

At supine position, the average heart rate was 76.9 (±13.4) beats per minute, and Bazett QTc interval was 428ms (±26.8ms); 428ms (±27.2ms) for female; 432ms (±23.8ms) for male). Ten patients, six females and four males, had prolonged QTc intervals (>460ms for women and >450ms for men (Giudicessi et al., 2019)) at baseline.

Response rates were 32.7%, and 37.0% at six- and 12-months follow-up, respectively. Of note, two patients had low baseline MADRS scores (10 and 8, respectively) and both exhibited a fluctuating symptom course over the treatment period.

### Baseline QTc lengths and MADRS change

Pearson’s correlation analysis revealed a significant association between QTc length and MADRS-score reduction, at six-months (r = -.34, p = .026), but not at 12-months follow-up (r = -.20, p = .208). Taking age, sex, BMI, and QTc prolongation upon risk medication into account in a linear regression analysis, QTc did not predict MADRS score reduction after six- (R^2^=.14, F=1.37, p=.330) nor at 12-months (R^2^=.10, F=.96, p=.444) of VNS treatment.

### Blood-based markers and depressive symptoms score

Exploratory correlation analyses after 12 months yielded that improvements of depressive symptoms were more pronounced in patients with higher MADRS scores as baseline (p < .001), lower ANC (p = .020), lower NLR (p = .017), and lower SII (p = .017; see table 2).

**Table 2.** Exploratory analyses of correlations between changes in MADRS-scores after six-, and 12-months VNS and sociodemographic, clinical, and blood-based markers in 22 patients.

|  | 6 months follow-up |  |  | 12 months follow-up |  |  |
| --- | --- | --- | --- | --- | --- | --- |
|  | Pearson's r | t | p | Pearson's r | t | p |
| Age (baseline) | -0.036 |  | 0.807 | -0.038 |  | 0.802 |
| Sex (female vs male) |  | 1.430 | 0.162 |  | 1.717 | 0.094 |
| BMI (baseline) | -0.072 |  | 0.649 | -0.017 |  | 0.915 |
| MADRS baseline | 0.524 |  | **<0.001** | 0.615 |  | **<0.001** |
| ANC | -0.451 |  | 0.060 | -0.527 |  | **0.020** |
| Lymphocyte count | 0.273 |  | 0.259 | 0.255 |  | 0.278 |
| Basophil count | 0.154 |  | 0.528 | 0.207 |  | 0.528 |
| Eosinophil count | 0.277 |  | 0.349 | 0.277 |  | 0.382 |
| Monocyte count | -0.043 |  | 0.863 | -0.224 |  | 0.344 |
| Platelet count | -0.128 |  | 0.601 | -0.165 |  | 0.488 |
| NLR | -0.430 |  | 0.075 | -0.540 |  | **0.017** |
| MLR | -0.175 |  | 0.473 | -0.258 |  | 0.273 |
| PLR | -0.230 |  | 0.344 | -0.235 |  | 0.318 |
| SII | -0.432 |  | 0.074 | -0.540 |  | **0.017** |
| Glucose | -0.026 |  | 0.915 | -0.005 |  | 0.982 |
| TG | 0.192 |  | 0.461 | 0.123 |  | 0.626 |
| HDL | 0.079 |  | 0.764 | 0.179 |  | 0.508 |
| LDL | 0.291 |  | 0.258 | 0.252 |  | 0.346 |
| CHOL | 0.244 |  | 0.345 | 0.134 |  | 0.597 |

Note: ANC = absolute neutrophil count; BMI = body mass index; CHOL = total cholesterol; HDL = high-density-lipoprotein; HR = heart rate; LDL = low-density lipoprotein; MADRS = Montgomery-Åsberg Depression Rating Scale; MLR = monocyte to lymphocyte ratio; NLR = neutrophil to lymphocyte ratio; PLR = platelet to lymphocyte ratio; SII = systemic inflammatory index

Given the exploratory nature of the analysis and the limited sample size relative to the number of variables we generated a stepwise regression with a forward selection approach to determine the main contributions of blood-based and anthropometric markers to MADRS scores reduction. The model for MADRS score reduction after six-months was significant (R^2^=.66, F=5.28, p=.013, see table 3). Here, baseline lower ANC, lower BMI, younger age and higher LDL concentrations correlated with MADRS reduction after six-months treatment with VNS. We did not retain SII in final model, its contribution was largely captured by ANC, which the stepwise procedure selected as the stronger independent predictor. Only, baseline lower ANC (R^2^=.28, F=6.55, p=.020) correlated with MADRS reduction at 12 months.

**Table 3.**
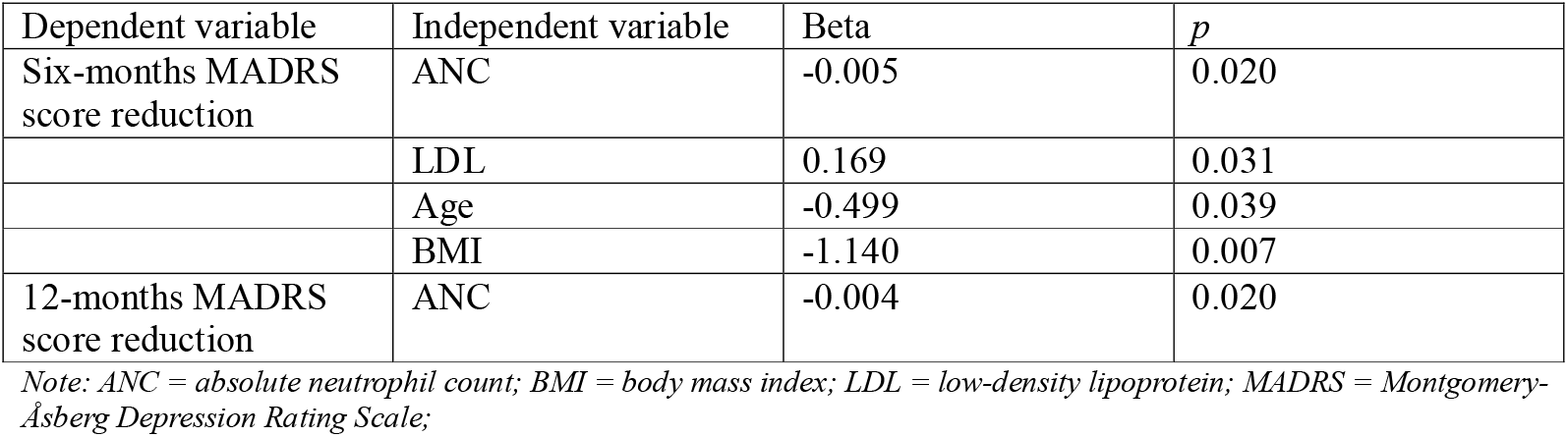
Stepwise multiple regression models for MADRS score reduction with hematological and sociodemographic variables as predictors.

## Discussion

In this retrospective data analysis, we tested the association between baseline QTc interval and depressive symptom reduction in patients suffering from DTD who received VNS adjunctive to treatment as usual. In contrast to a previously published result by Longpré-Poirier et al., in our sample baseline QTc time did not correlate with response to antidepressant VNS treatment. Beyond that, in exploratory analyses, we found four baseline variables potentially associated with later VNS response: absolute neutrophil count, BMI, age, and LDL.

Longpré-Poirier et al. (2020) reported that increased QTc intervals at baseline correlated with a greater reduction in depressive symptoms after 12 and 24-months antidepressant VNS measured with the HAM-D rating scale (Longpré-Poirier et al., 2020). Only slightly different items are captured in HAM-D compared to MADRS, which was the only available outcome scale at our centers. Both scales are highly positive correlated, so that we do not assume that different scales account for the difference of results between the two studies. Also, average QTc-times were similar in both studies (425.53+/-22 ms) While sample size was larger in our study pooled between three centers, we adjusted for relevant covariates such as sex, age, BMI, QTc prolongation by medication and baseline severity. Given the necessity to correct for sex differences in QTc analyses, we argue that this might largely account for differential findings. Of note, uncorrected results in our sample showed the opposite direction of association, highlighting the importance of proper adjustment.

Still, there is growing evidence that depressive symptomatology is associated with dysfunctions in the autonomic nervous system (Jandackova et al., 2016; Moretta & Messerotti Benvenuti, 2022; Watanabe et al., 2024). For instance, altered heart rate variability, a marker of autonomic function, has been associated with a vulnerability to depression independent from treatment with antidepressant medication (Jandackova et al., 2016; Moretta & Messerotti Benvenuti, 2022). More direct marker of the autonomic nervous system, such as the heart-rate variability or skin-conductance might serve as better baseline predictor. A study by authors of this study found that both in control subjects as well as in patients with depression VNS improved stress response only in subjects with lower baseline heart-rate variability (Schiweck et al., 2025).

In exploratory analyses we observed an association between ANC, BMI, and LDL with greater improvement in depressive symptoms severity. Due to limited sample size in a subsample of patients only and lack of correction for multiple testing, these results should be interpreted cautiously. Specifically, we found that ANC might be associated with greater improvement in depressive symptoms at six and 12 months after starting VNS. However, the near-zero Beta indicates that although statistically significant ANC had very limited practical impact. Elevated ANC has been associated with increased risk for depression, while lower ANC has been linked to greater symptom improvement during treatment in MDD (Elbakary et al., 2025; Foley et al., 2023). However, markers of low-grade inflammation such as NLR, MLR, PLR, and SII, that have been suggested as markers involved in the pathogenesis and the course of depression were not significant in our sample (Chen et al., 2025; Elbakary et al., 2025). Prior studies demonstrated an effect of VNS on the inflammatory system in MDD (Corcoran et al., 2005; Kavakbasi et al., 2024; Lespérance et al., 2024). However, these studies did not relate inflammatory changes to clinical symptom improvement. While inflammation may be involved in the pathogenesis of MDD in some cases, it might also be a response marker for antidepressant VNS treatment worth exploring .

In addition, we found an association between higher LDL levels at baseline and improvement of depressive symptoms severity following VNS. Notably, higher baseline LDL levels have been associated with greater symptom reduction in a study with mixed antidepressants (Wagner et al., 2019). Also, studies demonstrated alterations of lipid profiles after ECT or pharmacotherapy (Aksay et al., 2016; Gabriel, 2007; Hummel et al., 2011). In line with this literature, our findings suggest that baseline metabolic parameters such as LDL may be related to VNS outcomes. However, caution is warranted given above mentioned limitations in exploratory analyses.

In our sample, lower BMI was associated with improvement of depressive symptoms following VNS. Increased BMI has been consistently associated with MDD and treatment resistance (Kraus et al., 2023; Nour et al., 2023; Siau et al., 2025; L. Zhang et al., 2024). (Karageorgiou et al., 2023) In the present study, most individuals were overweight or obese, reflecting the high prevalence of metabolic dysregulation in DTD populations (Milaneschi et al., 2019). Results on the relationship between VNS and BMI in depression are limited (Kansagra et al., 2010) and warrants further investigations. Interestingly, VNS has also been explored in weight management and has received DFA approval for obesity treatment, underscoring the metabolic effects of vagal modulation.

Further, age was a significant marker of six-month depressive symptom severity, with higher age being associated with less symptom improvement. However, findings are inconsistent: a recent report from the RESTORE-LIFE study indicated that older age (>65 years) was associated with greater likelihood of response (Kavakbasi et al., 2025).

A small number of patients exhibited low baseline MADRS scores at study entry, including individuals meeting criteria for remission at the time of implantation. This reflects the real-world, multicenter nature of the study and clinical characteristics of patients with DTD, who often experience fluctuations of symptoms and unstable remission with high risk of relapse. All patients fulfilled study inclusion criteria and noMADRS cutoff was needed. In addition, response rates to VNS in our sample were smaller compared to previous investigations from other sites (Lespérance et al., 2024; Longpré-Poirier et al., 2020; Pigato et al., 2023; X. Zhang et al., 2022). However, our response rate coincides with reports from the RECOVER-trial (Rush et al., 2024).

### Limitations

There are several limitations in the presented work. For the exploratory analysis of blood-based makers data not from all centers were available. Thus, a better powered and controlled analysis of predictive markers would be needed to replicate exploratory analyses. We are aware of high chances of type-II error, but still presented these results, since data on variables correlating with antidepressant VNS are rare. Also, we did not have a control group nor ECGs at time-points after VNS. Therefore, longitudinal effects of VNS on QTc intervals cannot be evaluated. Additionally, in the study physical activity and smoking which were found to alter immune cell counts (Guo et al., 2024) was not evaluated. We measured serum lipids, and serum glucose, however, the study did not record medication intake for elevated TGs or hyperglycemia (Meng et al., 2025).

### Conclusion

With a larger sample and adjusting for relevant covariates, we did not replicate previous correlations between QTc intervals and depression score changes during VNS. However, exploratoryfindings suggest that baseline immune and metabolic variablesmight be associated with symptom improvements after VNS in DTD. Given the methodological limitations of a small sample size and lack of control group, these results should be interpreted with caution. Prospective and controlled studies may clarify whether baseline variables allow better selection of patients with higher chances of treatment response.

## Data availability

Data are available from the corresponding author upon reasonable request.

